# Towards Understanding Bipolar Disorder Through Social Media and Transformer Models: Challenges and Insights

**DOI:** 10.1101/2025.03.05.25323466

**Authors:** Vineet Srivastava, Lokesh Boggavarapu, Anthony Shin, Avisek Datta, Yingda Lu, Runa Bhaumik

## Abstract

Social media presents a promising avenue for monitoring mental health, yet detecting bipolar disorder (BD) remains significantly underexplored. The complexity arises from the overlap of linguistic patterns associated with depression and anxiety, making accurate identification challenging. This study aims to benchmark the performance of various transformer-based large language models (LLMs) trained on Reddit posts, to distinguish BD from other mental health conditions. Using a high-performing LLM (GPT-4o) as a benchmark, the analysis reveals that certain fine-tuned open small models (ex. MISTRAL, LLAMA) excel in capturing subtle linguistic cues linked to BD, achieving an F1 score of up to 0.86 with high precision and recall. However, BD was frequently misclassified as depression (range: 23%,51%), normal (range: 2%, 41%), and anxiety (range:1%,7%), underscoring the need for improved approaches. Integrating large language model insights with clinical expertise could significantly enhance bipolar disorder diagnosis and management.

## Introduction

According to an NIH report, 57.8 million adults live with some form of mental illness^1^. The rates of past year prevalence of mental illnesses in U.S. adults for anxiety disorders: 19.1%^2^, major depression: 8.3%^3^, and bipolar disorder: 2.8%^4^. Mental health problems affect a lot of people and create serious challenges, but research in this area often does not have the solid data that many other areas of healthcare do. This is partly because mental health issues are complex and hard to measure and partly because the stigma around mental illness has made it a difficult topic to openly discuss for a long time. Previous research showed that there is a significant symptom overlap between bipolar disorder and other mental health conditions like depression and anxiety ^5 6^. For instance, the presence of comorbid anxiety disorders in patients with BD has several adverse consequences, including a negative impact on almost all aspects of the presentation and course of BD^7–9^. Similarly, depressive episodes in bipolar disorder can resemble major depressive disorder, making it challenging to distinguish without considering the full spectrum of symptoms^10–14^.

Studying mental health often requires observing individuals’ behaviors—how they act, communicate, and interact with their surroundings, including relationships with friends and family. Traditional approaches primarily rely on periodic clinical evaluations and self-reported information, which can be subjective and may overlook subtle or temporary changes in mental states^15^. These limitations can lead to delays in diagnosis and intervention, potentially aggravating conditions before patients go to doctor’s offices^16^. For individuals with severe mental health conditions, the social network offers a place where people can fight stigma, feel empowered, and give hope to others through supportive online communities^17^ as well as to detect emotions, recognize psychological status, and predict mental conditions^18–22^.

Natural language processing (NLP), a subfield of artificial intelligence (AI) involves the ability to process, understand, and generate human language in a way that is meaningful. NLP heavily relies on machine learning (ML) techniques (especially deep learning models) to build models that can understand and generate natural language. The increasing use of natural language processing (NLP) and machine learning (ML) has significantly accelerated the study of mental health^23–28^ using clinical notes. Transformer-based LLMs have also shown significant advances in natural language processing tasks. Several pre-trained language models (PLMs), including BERT^29^, DistilBERT ^30^, ALBERT ^31^, BioClinical BERT^32^ for clinical notes, XLNET ^33^, and GPT model^34^ have been explored in the medical domain. In mental health studies, PLMs such as MentalBERT^35^, Mental-RoBERTa^35^, mental-xlnet-base-cased^36^ were developed to identify stress, anxiety, depression, and suicide. These studies demonstrated that employing pre-trained language representations in the target domain improved model performance on mental health detection tasks.

Numerous research has been found in diagnosing bipolar disorder from social media platforms^37^. For example, Linguistic and phonological features combined with a supervised machine-learning approach have been utilized to detect early signs of bipolar disorder from Twitter datal^38^. Similarly, pathway analysis has been applied to identify predictors of suicidal ideation in older adults with bipolar disorder by leveraging socio-demographically targeted advertisements on Facebook and newsfeeds^39^. Additionally, language impairments on Reddit have been examined to explore discussions in online mental health communities related to depression, bipolar disorder, and schizophrenia, focusing on expressions of positive emotions, exercise habits, and weight management^40^. Several machine learning algorithms, including Naïve Bayes^41^, convolutional neural network^42^, and Random Forest classifier^43^, have been employed to distinguish bipolar disorder from other categories through binary classification tasks on Reddit posts authored by users. Very little research has explored the use of fine-tuned transformer models in bipolar detection tasks. Other studies include transformer models as classification tasks utilizing ROBERTa^44^ and BERT-ATTN^45^. Further, a decision-support system was proposed using GPT-based large language models for BD treatment, highlighting the growing intersection of artificial intelligence and psychiatric care^46^.

To our knowledge, this is the first work that harnesses the capabilities of various advanced transformer-based algorithms fine-tuned with Reddit posts to solve a multiclassification task. Our study explores how well transformer models can identify bipolar disorder from social media posts, despite challenges like limited resources and similar symptoms in other mental health conditions.

Our research aims to answer the questions below:

1. How effectively can transformer-based language models identify bipolar disorder from social media posts?
2. How can bipolar disorder be distinguished in a multi-class prediction model?
3. What are the keywords that contribute to classification?
4. Are there any models comparable to the GPT series that perform well in low-resource settings?

## Methods

### Ethical Considerations

Reddit users are informed through Reddit’s Terms and Conditions that their posts are publicly accessible. In this study, no personally identifying information (e.g., names, locations, or IP addresses) was collected. Additionally, the authors did not engage in any discussions on the platform, eliminating the need to inform users that their posts might be used for research purposes. The data sets used in this study were obtained from different authors, as mentioned below, following a review by the University of Illinois Human Subjects Office. The submission was determined to fall under the category of “IRB-Exempt”.

### Data Collection

Reddit is a registered online platform that aggregates social news and facilitates online discussions. It is organized into various topic categories, with each specific area of interest referred to as a subreddit. The subreddit “Suicide Watch” (SW)^47^ serves as a key source of positive samples for annotation due to its focus on discussions related to suicide. In contrast, posts without suicidal content are collected from other popular subreddits to provide negative samples. The three datasets we combined are described below.

a. SWMH (SuicideWatch and Mental Health): Ji et al.^48^ collected this dataset from some mental health-related subreddits on Reddit sites to study mental disorders and suicidal ideation. This dataset contains discussions comprised of suicide-related intentions and mental disorders like depression, anxiety, and bipolar. Reddit’s official API was used, and a web spider was developed to collect the targeted forums. This collection contains 54,412 posts.
b. C-SSRS (Columbia Suicide Severity Rating Scale): Gaur et al.^49^ developed a Reddit C-SSRS Suicide Dataset, which consists of 500 user posts. These posts are labelled by four psychiatrists on a five-point scale according to guidelines established in the Columbia Suicide Severity Rating Scale, which progresses according to the severity of depression. As this dataset is clinically verified and labelled, it is an adequate dataset to validate the model. The posts were annotated in five different classes: supportive, suicide indicator, suicidal ideation, suicidal behavior, and suicide attempt High standards in annotation were maintained with substantial inter-rater agreement of 0.76.
c. RSDD (Reddit Self-reported Depression Diagnosis dataset)^50^: This dataset consists of Reddit posts for approximately 9,000 users who have claimed to have been diagnosed with depression (“diagnosed users”) and approximately 107,000 matched control users. All posts made to mental health-related subreddits or containing keywords related to depression were removed from the diagnosed users’ data; control users’ data do not contain such posts due to the selection process.

We considered five types of mental states: depression, suicide watch, anxiety, bipolar, and Normal from Reddit posts. As suicidal and depressed individuals often share similar patterns of language use, reflecting the overlapping cognitive and emotional states associated with these conditions, we have combined depression and suicide watch in one category. Table 1 shows the distribution of the annotated posts in each category for fine-tuning and evaluating the model. It is worth noting that this dataset is imbalanced due to the posts collected by the provider.

**Table 1:** Distribution of posts across training and evaluation dataset.

| Mental Disorder | Training Dataset | Evaluation Dataset | Total |
| --- | --- | --- | --- |
|  | N (%) | N (%) |  |
| Depression/SuicideWatch | 24276 (50%) | 4646 (47%) | 28922 |
| Normal | 10000 (20%) | 2500 (25%) | 12500 |
| Anxiety | 8042 (17%) | 1508 (16%) | 9550 |
| Bipolar | 6408 (13%) | 1220 (12%) | 7628 |
| Total | 48726 | 9874 | 58600 |

Note: The training dataset was divided into 80% for training and 20% for testing during fine-tuning process.

### Feature Engineering

To classify text using a machine learning or deep learning approach, feature engineering methods have been applied to convert text data into numerical values. Feature engineering for advanced language models like ALBERT, MentalBERT, BioClinicalBERT, LLAMA, Mistral, and GPT-4 primarily focused on efficient tokenization, input representation, and preprocessing rather than traditional manual feature extraction. These models relied on pre-trained embeddings and sophisticated tokenization techniques to handle diverse and complex data inputs. For instance, ALBERT and BioClinicalBERT used WordPiece tokenization, with the latter optimized for domain-specific tasks in biomedical and clinical contexts. These models preprocessed input by normalizing text, handling domain-specific jargon, and formatting input sequences with special tokens like [CLS] and [SEP] to represent sentence relationships.

LLAMA and Mistral employed more advanced tokenization methods like SentencePiece or Byte Pair Encoding (BPE), which were designed to process multilingual and large-scale datasets efficiently. GPT-4 extended beyond text by integrating multi-modal input capabilities and dynamically formatting prompts to handle textual and non-textual data. Preprocessing for these models involved chunking long contexts, standardizing inputs, and leveraging embeddings for cross-modal tasks. For the machine learning model, we used TF-IDF (Term Frequency-Inverse Document Frequency) embedding to represent textual features.

### Language Models

#### ALBERT^51^

We utilized the ALBERT model and fine-tuned it with the training dataset to build the classifier. For implementation, we employed the Hugging Face Transformers library, an open-source library and data science platform that provides tools to build, train and deploy ML models. Text inputs were truncated to 128 tokens, and the models were trained with the following hyperparameters: learning_rate= 2e-5, power = 0.5, decay_steps =12, and epochs =12. The model was trained using the AdamW optimizer^52^. The best-performing model was selected based on the evaluation of the weighted F1 score on a validation dataset. A random 20% split of the fine-tuned data, without stratification, was reserved for testing the model during the fine-tuning process. Fine-tuning of the model was conducted using Amazon SageMaker, a comprehensive cloud platform that facilitates the development, training, and deployment of machine learning models at scale. This platform provided the necessary computational resources and tools to optimize the model’s performance for specific tasks.

#### MentalBERT^35^

We utilized a BERT model pre-trained on mental health-related texts and fine-tuned it for mental health classification tasks. The model was implemented using the Huggingface Transformers library, which provides pre-trained language models and tools for natural language processing tasks. The dataset was split into training (80%), validation (10%), and test (10%) sets, comprising 43,853, 4,873, and 9,874 samples respectively. The model was trained to classify texts into four categories: Depression/suicideWatch, Anxiety, Normal, and Bipolar. Input texts were tokenized using the model’s native tokenizer and standardized to a maximum length of 128 tokens. The fine-tuning process employed the following hyperparameters: learning rate of 2e-5, batch size of 8 for both training and evaluation, weight decay of 0.01, and 4 training epochs. We used the AutoModelForSequenceClassification framework for multi-label classification. The training implemented an epoch-based evaluation strategy, saving the best-performing model based on validation set performance.

#### BioClinical BERT^53^

We employed a domain-adapted BERT model pre-trained on clinical texts and fine-tuned it for mental health classification tasks. The implementation utilized the Huggingface Transformers library for model architecture and training procedures. The model was trained on the same dataset split as MentalBERT. Fine-tuning was conducted using identical hyperparameters to ensure fair comparison: learning rate of 2e-5, batch size of 8 for both training and evaluation, weight decay of 0.01, and 4 training epochs. The model utilized the AutoModelForSequenceClassification framework for multi-label classification. Training progress was evaluated after each epoch, with model checkpoints saved based on validation performance. Both models’ performance was evaluated using precision, recall, and F1-score metrics, calculated for individual classes and as weighted averages. Confusion matrices were generated to visualize the classification performance across all categories.

#### GPT-4o^34^

We utilized OpenAI’s GPT-4o model, an advanced Generative Pre-trained Transformer (GPT). We set temperature parameter to .5 and a fixed random seed. GPT-4o (like other OpenAI models) is inherently non-deterministic by default, meaning different runs can yield slightly different outputs even for the same prompt.

#### LLAMA^54^

(Large Language Model Meta AI): We fine-tuned a LLaMA-3.2 model using QLoRA, optimizing it for mental health classification tasks. The implementation leveraged the Hugging Face transformers library and PEFT for parameter-efficient fine-tuning. The model was trained with a batch size of 1 per device and a gradient accumulation step of 8 to effectively utilize memory resources. The training utilized the Paged AdamW optimizer with a learning rate of 2e-4, weight decay of 0.001, and a cosine learning rate scheduler with a warmup ratio of 0.03. Evaluation metrics included precision, recall, and F1-score, computed both per class and as weighted averages. Confusion matrices were generated to provide insights into classification performance across different categories. The training process and results were logged using Weights & Biases (W&B) for better tracking and visualization.

#### Mistral^55^

We fine-tuned the Mistral-7B model using QLoRA for mental health classification tasks. To ensure a fair comparison, the training setup and hyperparameters were kept identical to those used for LLAMA fine-tuning. Key parameters, including LoRA configuration, learning rate, batch size, optimizer, gradient checkpointing, and learning rate scheduling, remained consistent. Model performance was assessed using confusion matrices like other models.

### Machine Learning Algorithm

We used Random Forest^56^ algorithm as a baseline approach for multi-class classification. The parameter grid for hyperparameter tuning was set with the following values: ‘n_estimators’ to [100, 200, 300], ‘max_depth’ to [10], ‘min_samples_split’ to [2, 5], and ‘min_samples_leaf’ to [1,2].

## Results

The word cloud visualizations in Figure 1 highlight the common themes and overlaps between the depression and SuicideWatch categories. Based on the relative frequency and prominent words depicted in each word cloud, both categories share several high-frequency words, which highlight the overlap in language used to express emotions related to depression and suicidal thoughts. Common high-frequency words include: “feel,” “time,” “want,” “know,” and “thing.” In depression category, words like “depressed,” “depression,” “sad,” “hate,” and “alone” are prominent. These words suggest a focus on the enduring emotional state and feelings of isolation and negativity. In the SuicideWatch category, words such as “die,” “suicide,” “kill,” and “end” stand out. This points to more direct expressions related to the act of suicide and finality, indicating acute distress and potential crisis situations. Given the significant overlap in language patterns observed between individuals expressing feelings of depression and those under suicide watch, it is evident that these conditions share common cognitive and emotional underpinnings.

**Figure 1.**
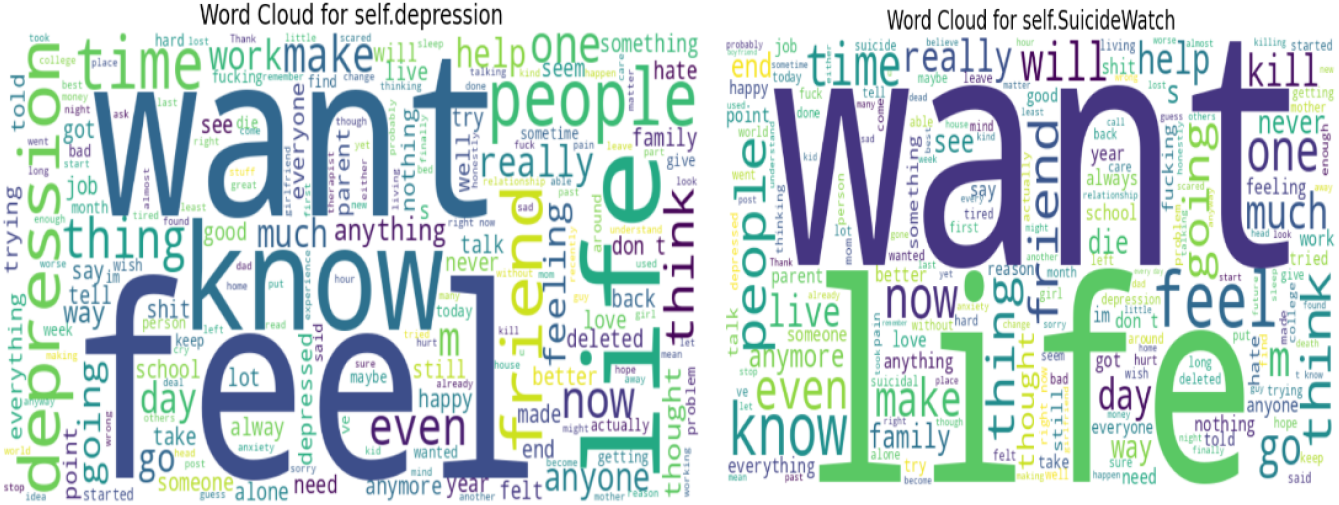
Word visualization of depression and SuicideWatch categories

### Performance Evaluation of Fine-tuned Models

The classification performance during the fine-tuning process (Table 2) reveals that transformer-based models MISTRAL7B (.85), LLAMA3.2(0.80), and MentalBERT (.73) outperform recall measures than the traditional Random Forest (TF-IDF: .57), highlighting the advantage of contextual understanding in text classification for bipolar posts

**Table 2:** Classification performances during the fine-tuning process for bipolar disorder.

| Models | F1 | Precision | Recall |
| --- | --- | --- | --- |
| ALBERT | .72 | .78 | .67 |
| MentalBERT | .79 | .88 | .73 |
| BioClinical BERT | .77 | .86 | .70 |
| LLAMA 3.2 | .83 | .87 | .80 |
| MISTRAL7B | .86 | .88 | .85 |
| RF (TF-IDF) | .70 | .90 | .57 |

### Performance Evaluation on Validation Dataset

The classification performance (Table 3) in the Bipolar category varies significantly across models (F1 measures ranging from .67 to .86). MISTRAL7B achieves the highest F1 score of 0.86, balancing strong precision (0.88) with recall (0.85). LLAMA 3.2 follows closely with an F1 score of 0.83, benefiting from high precision (0.87) and recall (0.80). ALBERT demonstrates weaker performance with an F1 score of 0.67, primarily due to lower recall (0.60). MentalBERT (F1= .78) and BioClinicalBERT (F1= .77) have similar accuracy. GPT-4o achieves an F1 score of 0.72, driven by excellent precision (0.96) but low recall (0.58). Traditional methods like TF-IDF + RF perform moderately well, with an F1 score of 0.70, balancing strong precision (0.91) and low recall (0.56). Consequently, high detection rates for depression/suicidewatch (.81-.94), normal (.72-.94), anxiety (.62-.83) across all models have been observed in supplementary Table 1.

**Table 3:** Classification performances on evaluation dataset for bipolar disorder posts.

| Models | F1 | Precision | Recall |
| --- | --- | --- | --- |
| ALBERT | 0.67 | 0.77 | 0.6 |
| MentalBERT | 0.78 | 0.86 | 0.72 |
| BioClinical BERT | 0.77 | 0.85 | .70 |
| GPT-4o | 0.72 | 0.96 | 0.58 |
| LLAMA 3.2 | 0.83 | 0.87 | 0.80 |
| MISTRAL7B | 0.86 | 0.88 | 0.85 |
| RF (TF-IDF) | 0.69 | 0.89 | 0.56 |

### Misclassification Analysis

Figure 2 shows the misclassification percentages of bipolar disorder into three categories: Anxiety, Depression, and Normal, across different models. Depression is the most frequent misclassification across all models, with the highest percentages ranging from 20% (MentalBERT) to 51% (LLAMA 3.2). Anxiety is less frequently misclassified compared to depression, with percentages ranging from 1% (LLAMA 3.2) to 7% (Albert and GPT-4o). Normal has generally low misclassification rates, except for LLAMA 3.2 (41%) and MISTRAL7b (14%) The results suggest that bipolar disorder has been misclassified as depression more frequently than as anxiety or normal categories.

**Figure 2:**
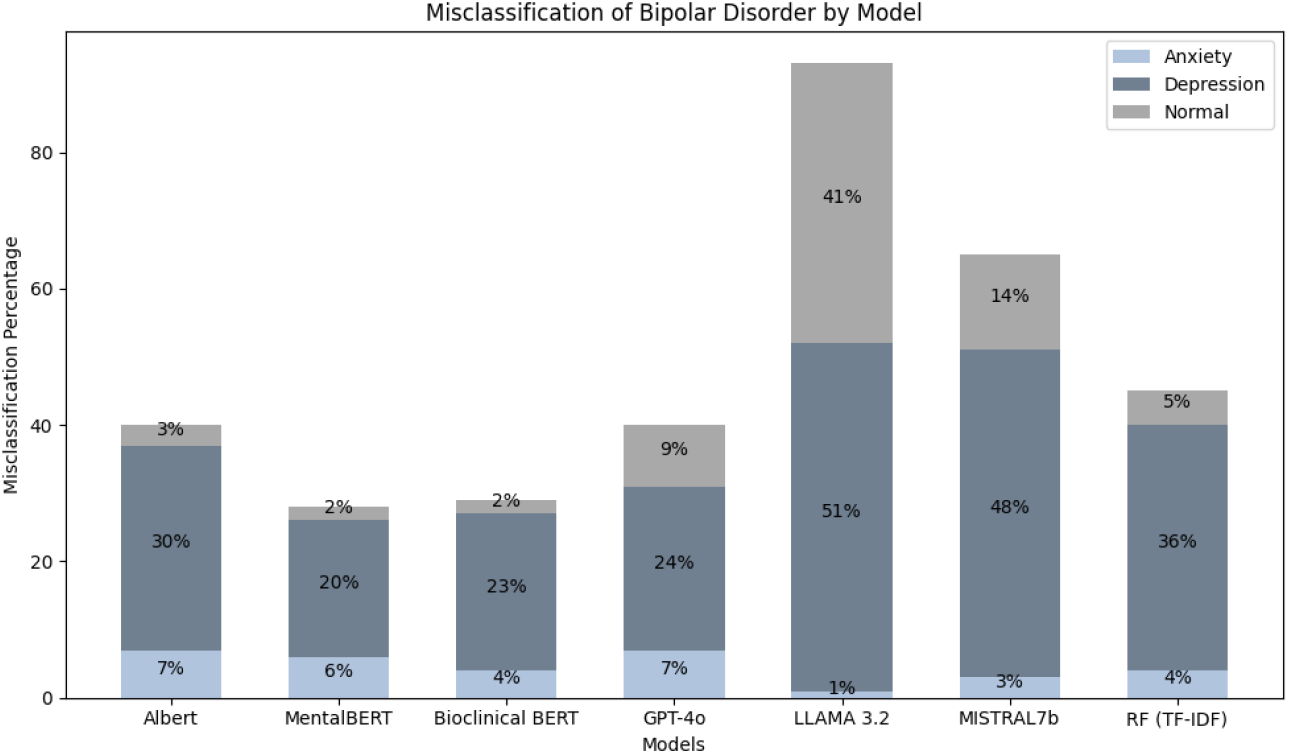
Misclassification Analysis for Bipolar Disorder

### Model Interpretability

We used “Local Interpretable Model-agnostic Explanations (LIME)” designed to improve the transparency and interpretability of complex deep-learning models. LIME highlights which features are important for a specific prediction, enabling users to focus on the most relevant inputs. For the random forest algorithm, we used the Gini impurity^57^ for the feature importance score. Figure 3 displays the importance of keywords for four different models.

**Figure 3:**
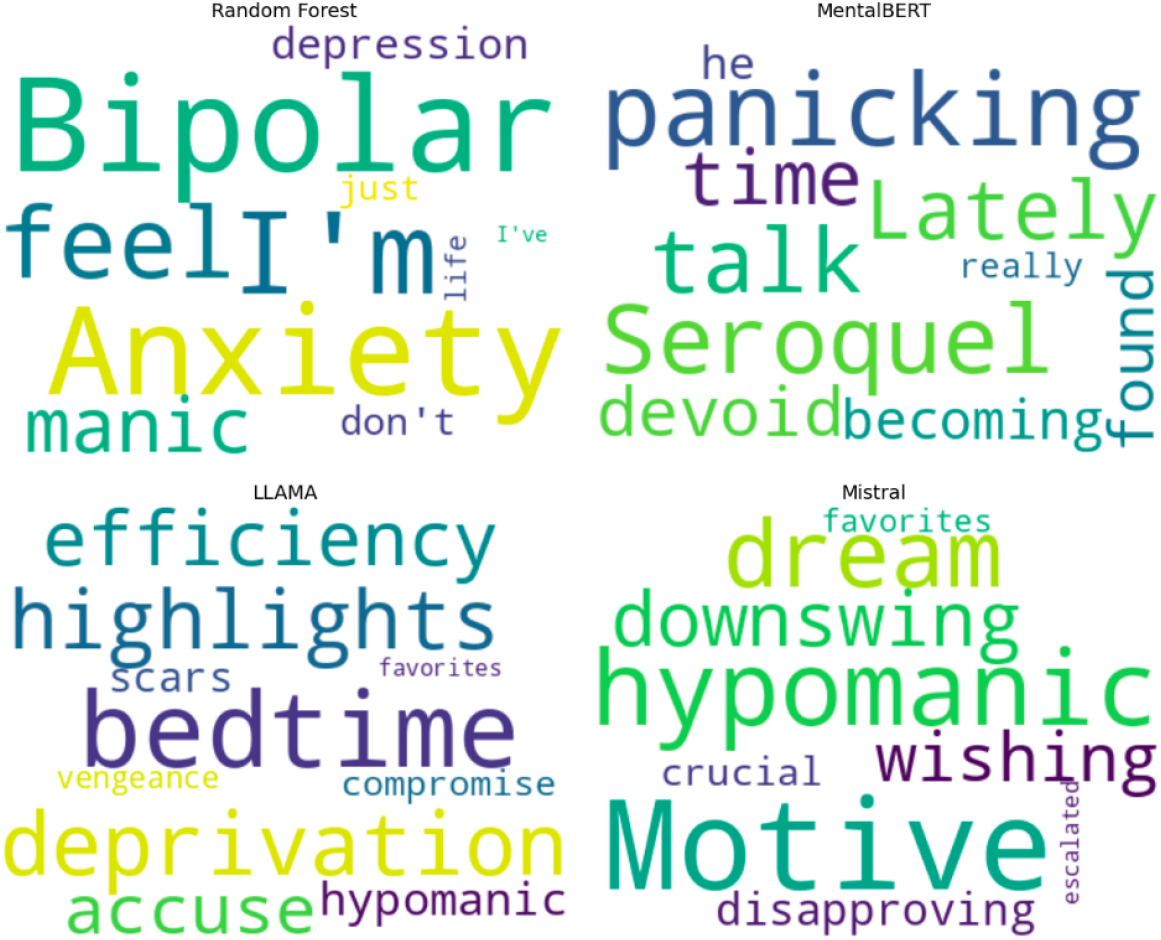
Top words visualization for bipolar disorder in four LLMs.

The words identified (e.g., bipolar, anxiety, manic, depression, meds, episode, feel) in TF-IDF seem frequent in the dataset but also carry high importance in distinguishing between different classes. They are commonly used in discussions related to bipolar disorder. Moreover I’m” and “I’ve” indicated a high frequency of self-referential language, which is common in BD, especially during manic or depressive episodes. MentalBERT selects words like “panicking”, “Seroquel”, “devoid”, “found”, “becoming”, and “talk”, indicating that the model captures medical terms and emotional states more effectively than TF-IDF as it is specifically fine-tuned on mental health-related texts. For example, “Seroquel” is a brand name for quetiapine, an atypical antipsychotic medication commonly prescribed to manage symptoms of BD, including manic and depressive episodes. While not exclusive to BD, experiences of panic or anxiety can occur during depressive or mixed episodes in individuals with BD. These terms are not specifically linked to BD but could appear in personal narratives or discussions related to the disorder. For example, “talk” might refer to therapy sessions, and “becoming” could relate to changes in mood or behavior. However, these words are common in various contexts and are not unique indicators of BD.

LLAMA suggests terms such as “bedtime”, “deprivation”, “efficiency”, “hypomanic”, and “vengeance” that focuses on sleep-related patterns^58^, energy levels, and behavioral descriptions rather than direct medical terms. As LLAMA is not specialized in mental health but instead generalizes from a broad set of data, it may lead to identifying words that are linguistically rich but not necessarily medical or clinical terms. Similar results are shown for MISTRAL7B, where words like “hypomanic”, “downswing”, “motive” and “escalated” are linked to the bipolar disorder condition^59^. Evidently, large language models selected subtle linguistic cues that appear in mental health discussions in social media posts.

## Discussions

This study assessed the performance of several models, including GPT-4o, ALBERT, mentalBERT, BioClinicalBERT, Random Forest (utilizing both tf-idf and BERT embeddings), LLAMA 3.2, and MISTRAL7B, across four mental health classification tasks: depression/suicide watch, normal, anxiety, and bipolar disorder. Notably, we emphasized bipolar disorder, an underexplored area within mental health research.

The classification of bipolar disorder posed challenges across models, highlighting the inherent difficulty in distinguishing its patterns effectively. While GPT-4o exhibited strong precision (.96), it struggled with recall (.58), indicating that they were conservative in identifying bipolar cases, likely to avoid false positives. Others such as LLAMA 3.2 (.80) and MISTRAL7B (.85) demonstrated higher recall but at the cost of few false positives Both models are most effective, providing a more balanced performance by successfully identifying a greater number of bipolar cases without compromising precision. Previous studies have approached the identification of bipolar disorder using binary classification methods. For instance, a Naive Bayes classifier utilizing n-gram features achieved a detection rate of 60% for bipolar disorder^41^, compared to detection rates nearing 90% for other mental health disorders. Similarly, a convolutional neural network^42^ (CNN) identified bipolar disorder with a detection rate of only 38%. In contrast, the ROBERTa^44^ language model demonstrated a recall rate of 79%, while the Random Forest classifier^43^ attained an impressive F1 measure of 0.86. Studies showed that in a multiclass classification approach, BERT-ATTN^45^ with logistic regression achieves an F1 score of .879, and with multilabel and multi-class classification models^60^, it achieves an F1 score between 34.58 and 51.64. In this study, LLAMA 3.2, and MISTRAL7B achieved higher performance than previous studies in bipolar detection. Further, it was observed that people with bipolar disorder were much more likely to use words like “I” and “me” compared to Reddit users who didn’t report having the condition^60^. They connected this pattern to “authenticity”, meaning people with bipolar disorder may express themselves more honestly.

Our study has several limitations that should be acknowledged. First, Reddit posts are self-reported and lack clinical validation, making it difficult to directly apply findings to real-world clinical settings. These posts may include irrelevant or misleading information that could distort analysis. Second, we have merged depression and suicide watch into a single category due to the similarities in language use and the fact that depression is the primary contributor to suicide rates. However, it’s crucial to acknowledge the specific differences that could impact treatment and intervention approaches. Third, data imbalance may bias the traditional and language model toward the majority class, leading to inferior performance in underrepresented classes like Bipolar. Fourth, Social media users do not represent the entire bipolar population, and language patterns vary across platforms (Reddit vs. Twitter), affecting model generalizability. Finally, a longitudinal assessment of bipolar disorder posts was not considered.

To address these abovementioned limitations, future work will focus on several key areas. First, we will aim to incorporate clinically validated datasets or collaborate with mental health professionals to create a more reliable benchmark for real-world applicability. This will help bridge the gap between self-reported social media data and clinical relevance to reduce the risk of incorporating irrelevant or misleading information. Second, expanding the dataset to include a more diverse range of mental health conditions and user demographics will further improve the model’s robustness and generalizability. By building upon these directions, future work will aim to develop a more clinically relevant AI system for mental health analysis.

## Data Availability

All data produced in the present study are available upon reasonable request to the authors

## Data Availability

The data used to support the findings of this study are available from the corresponding author upon request.

## Code Availability

Code is available from the corresponding author upon request.

## Acknowledgements

The authors thank AI.Health4All Center and the Department of Psychiatry in the College of Medicine, University of Illinois Chicago, for their support in providing computing resources and research.

## Author Contributions

R.B. and V.S. conceptualized the problem and collected data. R.B., V.S., L.B., and A.S. contributed to the coding and method section. A.D. contributed to coding. Y.L. contributed valuable feedback on the manuscript. R.B. prepared the manuscript.

## References

1. National Institute of Mental Health. National Institute of Mental Health, “Mental Illness,.”; 2023. https://www.nimh.nih.gov/health/statistics/mental-illness

2. National Institute of Mental Health, “Any Anxiety Disorder,.” https://www.nimh.nih.gov/health/statistics/any-anxiety-disorder.

3. National Institute of Mental Health, “Major Depression,.” https://www.nimh.nih.gov/health/statistics/major-depression.

4. National Institute of Mental Health, “Bipolar Disorder,.” https://www.nimh.nih.gov/health/statistics/bipolar-disorder.

5. American PsychiatricAssociation. https://www.psychiatry.org/

6. Bauer MS, Altshuler L, Evans DR, Beresford T, Williford WO, Hauger R. Prevalence and distinct correlates of anxiety, substance, and combined comorbidity in a multi-site public sector sample with bipolar disorder. Journal of Affective Disorders. 2005;85(3):301–315. doi:10.1016/j.jad.2004.11.009

7. Spoorthy MS, Chakrabarti S, Grover S. Comorbidity of bipolar and anxiety disorders: An overview of trends in research. World J Psychiatry. 2019;9(1):7–29. doi:10.5498/wjp.v9.i1.7

8. Nabavi B, Mitchell AJ, Nutt D. A Lifetime Prevalence of Comorbidity Between Bipolar Affective Disorder and Anxiety Disorders: A Meta-analysis of 52 Interview-based Studies of Psychiatric Population. EBioMedicine. 2015;2(10):1405–1419. doi:10.1016/j.ebiom.2015.09.006

9. Sasson Y, Chopra M, Harrari E, Amitai K, Zohar J. Bipolar comorbidity: from diagnostic dilemmas to therapeutic challenge. Int J Neuropsychopharm. 2003;6(2):139–144. doi:10.1017/S1461145703003432

10. Angst J, Sellaro R, Stassen HH, Gamma A. Diagnostic conversion from depression to bipolar disorders: results of a long-term prospective study of hospital admissions. J Affect Disord. 2005;84(2-3):149–157. doi:10.1016/S0165-0327(03)00195-2

11. Goldberg JF, Harrow M, Whiteside JE. Risk for Bipolar Illness in Patients Initially Hospitalized for Unipolar Depression. AJP. 2001;158(8):1265–1270. doi:10.1176/appi.ajp.158.8.1265

12. Holma KM, Melartin TK, Holma IAK, Isometsä ET. Predictors for Switch From Unipolar Major Depressive Disorder to Bipolar Disorder Type I or II: A 5-Year Prospective Study. J Clin Psychiatry. 2008;69(8):1267–1275. doi:10.4088/JCP.v69n0809

13. Inoue T, Kimura T, Inagaki Y, Shirakawa O. Prevalence of Comorbid Anxiety Disorders and Their Associated Factors in Patients with Bipolar Disorder or Major Depressive Disorder. Neuropsychiatr Dis Treat. 2020;16:1695–1704. doi:10.2147/NDT.S246294

14. Goes FS, McCusker MG, Bienvenu OJ, et al. Co-morbid anxiety disorders in bipolar disorder and major depression: familial aggregation and clinical characteristics of co-morbid panic disorder, social phobia, specific phobia and obsessive-compulsive disorder. Psychol Med. 2012;42(7):1449–1459. doi:10.1017/S0033291711002637

15. Naslund JA, Aschbrenner KA, Marsch LA, Bartels SJ. The future of mental health care: peer-to-peer support and social media. Epidemiol Psychiatr Sci. 2016;25(2):113–122. doi:10.1017/S2045796015001067

16. Moorhead SA, Hazlett DE, Harrison L, Carroll JK, Irwin A, Hoving C. A New Dimension of Health Care: Systematic Review of the Uses, Benefits, and Limitations of Social Media for Health Communication. J Med Internet Res. 2013;15(4):e85. doi:10.2196/jmir.1933

17. Coppersmith G, Leary R, Crutchley P, Fine A. Natural Language Processing of Social Media as Screening for Suicide Risk. Biomed Inform Insights. 2018;10:117822261879286. doi:10.1177/1178222618792860

18. Reavley NJ, Pilkington PD. Use of Twitter to monitor attitudes toward depression and schizophrenia: an exploratory study. PeerJ. 2014;2:e647. doi:10.7717/peerj.647

19. Nguyen T, Phung D, Dao B, Venkatesh S, Berk M. Affective and Content Analysis of Online Depression Communities. IEEE Trans Affective Comput. 2014;5(3):217–226. doi:10.1109/TAFFC.2014.2315623

20. Islam MdR, Kabir MA, Ahmed A, Kamal ARM, Wang H, Ulhaq A. Depression detection from social network data using machine learning techniques. Health Inf Sci Syst. 2018;6(1):8. doi:10.1007/s13755-018-0046-0

21. O’Dea B, Wan S, Batterham PJ, Calear AL, Paris C, Christensen H. Detecting suicidality on Twitter. Internet Interventions. 2015;2(2):183–188. doi:10.1016/j.invent.2015.03.005

22. Trotzek M, Koitka S, Friedrich CM. Utilizing Neural Networks and Linguistic Metadata for Early Detection of Depression Indications in Text Sequences. IEEE Trans Knowl Data Eng. 2020;32(3):588–601. doi:10.1109/TKDE.2018.2885515

23. Librenza-Garcia D, Kotzian BJ, Yang J, et al. The impact of machine learning techniques in the study of bipolar disorder: A systematic review. Neuroscience & Biobehavioral Reviews. 2017;80:538–554. doi:10.1016/j.neubiorev.2017.07.004

24. Monteith S, Glenn T, Geddes J, Whybrow PC, Bauer M. Big data for bipolar disorder. Int J Bipolar Disord. 2016;4(1):10. doi:10.1186/s40345-016-0051-7

25. Alonso SG, de la Torre-Díez I, Hamrioui S, et al. Data Mining Algorithms and Techniques in Mental Health: A Systematic Review. J Med Syst. 2018;42(9):161. doi:10.1007/s10916-018-1018-2

26. Calvo RA, Milne DN, Hussain MS, Christensen H. Natural language processing in mental health applications using non-clinical texts. Nat Lang Eng. 2017;23(5):649–685. doi:10.1017/S1351324916000383

27. Le Glaz A, Haralambous Y, Kim-Dufor DH, et al. Machine Learning and Natural Language Processing in Mental Health: Systematic Review. J Med Internet Res. 2021;23(5):e15708. doi:10.2196/15708

28. Harvey D, Lobban F, Rayson P, Warner A, Jones S. Natural Language Processing Methods and Bipolar Disorder: Scoping Review. JMIR Ment Health. 2022;9(4):e35928. doi:10.2196/35928

29. Eriksen AV, Möller S, Ryg J. Use of GPT-4 to Diagnose Complex Clinical Cases. NEJM AI. 2024;1(1). doi:10.1056/AIp2300031

30. Malviya K, Roy B, Saritha S. A Transformers Approach to Detect Depression in Social Media. In: 2021 International Conference on Artificial Intelligence and Smart Systems (ICAIS). IEEE; 2021:718–723. doi:10.1109/ICAIS50930.2021.9395943

31. Haque F, Nur RU, Jahan SA, Mahmud Z, Shah FM. A Transformer Based Approach To Detect Suicidal Ideation Using Pre-Trained Language Models. In: 2020 23rd International Conference on Computer and Information Technology (ICCIT). IEEE; 2020:1–5. doi:10.1109/ICCIT51783.2020.9392692

32. Kshatriya B, Nunez N, Gardea Resendez M, et al. Neural Language Models with Distant Supervision to Identify Major Depressive Disorder from Clinical Notes.; 2021.

33. Wang X, Chen S, Li T, et al. Depression Risk Prediction for Chinese Microblogs via Deep-Learning Methods: Content Analysis. JMIR Med Inform. 2020;8(7):e17958. doi:10.2196/17958

34. “OpenAI (2023). GPT-4 Technical Report.

35. Ji S, Zhang T, Ansari L, Fu J, Tiwari P, Cambria E. MentalBERT: Publicly Available Pretrained Language Models for Mental Healthcare. Published online 2021. doi:10.48550/ARXIV.2110.15621

36. Ji S, Zhang T, Yang K, Ananiadou S, Cambria E, Tiedemann J. Domain-specific Continued Pretraining of Language Models for Capturing Long Context in Mental Health. Published online 2023. doi:10.48550/ARXIV.2304.10447

37. Timakum T, Xie Q, Lee S. Identifying mental health discussion topic in social media community: subreddit of bipolar disorder analysis. Front Res Metr Anal. 2023;8:1243407. doi:10.3389/frma.2023.1243407

38. Huang YH, Wei LH, Chen YS. Detection of the Prodromal Phase of Bipolar Disorder from Psychological and Phonological Aspects in Social Media. Published online December 26, 2017. doi:10.48550/arXiv.1712.09183

39. O’Rourke N, Heisel MJ, Canham SL, Sixsmith A, BADAS Study Team. Predictors of suicide ideation among older adults with bipolar disorder. Hashimoto K, ed. PLoS ONE. 2017;12(11):e0187632. doi:10.1371/journal.pone.0187632

40. Park A, Conway M. Harnessing Reddit to Understand the Written-Communication Challenges Experienced by Individuals With Mental Health Disorders: Analysis of Texts From Mental Health Communities. J Med Internet Res. 2018;20(4):e121. doi:10.2196/jmir.8219

41. Gkotsis G, Oellrich A, Velupillai S, et al. Characterisation of mental health conditions in social media using Informed Deep Learning. Sci Rep. 2017;7(1):45141. doi:10.1038/srep45141

42. Kim Y. Convolutional Neural Networks for Sentence Classification. In: Proceedings of the 2014 Conference on Empirical Methods in Natural Language Processing (EMNLP). Association for Computational Linguistics; 2014:1746–1751. doi:10.3115/v1/D14-1181

43. Sekulic I, Gjurković M, Šnajder J. Not Just Depressed: Bipolar Disorder Prediction on Reddit. In: Proceedings of the 9th Workshop on Computational Approaches to Subjectivity, Sentiment and Social Media Analysis. Association for Computational Linguistics; 2018:72–78. doi:10.18653/v1/W18-6211

44. Murarka A, Radhakrishnan B, Ravichandran S. Detection and Classification of mental illnesses on social media using RoBERTa. Published online 2020. doi:10.48550/ARXIV.2011.11226

45. Jiang Z, Levitan SI, Zomick J, Hirschberg J. Detection of Mental Health from Reddit via Deep Contextualized Representations. In: Proceedings of the 11th International Workshop on Health Text Mining and Information Analysis. Association for Computational Linguistics; 2020:147–156. doi:10.18653/v1/2020.louhi-1.16

46. Perlis RH, Goldberg JF, Ostacher MJ, Schneck CD. Clinical decision support for bipolar depression using large language models. Neuropsychopharmacol. 2024;49(9):1412–1416. doi:10.1038/s41386-024-01841-2

47. Reddit. Reddit Suicide Watch Posts. https://www.reddit.com/r/SuicideWatch/

48. Ji S, Li X, Huang Z, Cambria E. Suicidal ideation and mental disorder detection with attentive relation networks. Neural Comput & Applic. 2022;34(13):10309–10319. doi:10.1007/s00521-021-06208-y

49. Gaur M, Alambo A, Sain JP, et al. Reddit C-SSRS Suicide Dataset. Published online May 4, 2019. doi:10.5281/ZENODO.2667859

50. Yates A, Cohan A, Goharian N. Depression and Self-Harm Risk Assessment in Online Forums. Published online 2017. doi:10.48550/ARXIV.1709.01848

51. Lan Z, Chen M, Goodman S, Gimpel K, Sharma P, Soricut R. ALBERT: A Lite BERT for Self-supervised Learning of Language Representations. Published online 2019. doi:10.48550/ARXIV.1909.11942

52. Loshchilov I, Hutter F. Decoupled Weight Decay Regularization. Published online January 4, 2019. doi:10.48550/arXiv.1711.05101

53. Alsentzer E, Murphy J, Boag W, et al. Publicly Available Clinical. In: Proceedings of the 2nd Clinical Natural Language Processing Workshop. Association for Computational Linguistics; 2019:72–78. doi:10.18653/v1/W19-1909

54. Touvron H, Lavril T, Izacard G, et al. LLaMA: Open and Efficient Foundation Language Models. Published online 2023. doi:10.48550/ARXIV.2302.13971

55. Mistral. https://mistral.ai/en

56. Breiman L. Random Forest. Machine Learning. 2001;45(1):5–32. doi:10.1023/A:1010933404324

57. Rokach L, Maimon O. Top-Down Induction of Decision Trees Classifiers—A Survey. IEEE Trans Syst, Man, Cybern C. 2005;35(4):476–487. doi:10.1109/TSMCC.2004.843247

58. Hensch T, Wozniak D, Spada J, et al. Vulnerability to bipolar disorder is linked to sleep and sleepiness. Transl Psychiatry. 2019;9(1):294. doi:10.1038/s41398-019-0632-1

59. Taylor E. Managing bipolar disorders in children and adolescents. Nat Rev Neurol. 2009;5(9):484–491. doi:10.1038/nrneurol.2009.117

60. Cohan A, Desmet B, Yates A, Soldaini L, MacAvaney S, Goharian N. SMHD: A Large-Scale Resource for Exploring Online Language Usage for Multiple Mental Health Conditions. Published online 2018. doi:10.48550/ARXIV.1806.05258

