## supplementary tables for "Towards Understanding Bipolar Disorder Through Social Media and Transformer Models: Challenges and Insights" (from another version of this preprint: version 1, 2025-03-06)

### Table 1: Classification performances of evaluation data

| Mental Disorder | Models | F1 | Precision | Recall |
|---|---|---|---|---|
| Depression/Suicide Watch | ALBERT | 0.81 | 0.81 | 0.81 |
| | MentalBERT | 0.86 | 0.81 | 0.92 |
| | BioClinical BERT | 0.88 | 0.85 | 0.92 |
| | GPT-4o | 0.85 | 0.83 | 0.89 |
| | LLAMA 3.2 | 0.90 | 0.87 | 0.93 |
| | MISTRAL7B | 0.92 | 0.90 | 0.94 |
| | RF (TF-IDF) | 0.82 | 0.73 | 0.92 |
| Normal | ALBERT | 0.76 | 0.7 | 0.83 |
| | MentalBERT | 0.88 | 0.90 | 0.87 |
| | BioClinical BERT | 0.86 | 0.1 | 0.82 |
| | GPT-4o | 0.8 | 0.80 | 0.80 |
| | LLAMA 3.2 | 0.92 | 0.94 | 0.90 |
| | MISTRAL7B | 0.95 | 0.96 | 0.94 |
| | RF (TF-IDF) | 0.78 | 0.84 | 0.72 |
| Anxiety | ALBERT | 0.73 | 0.78 | 0.69 |
| | MentalBERT | 0.79 | 0.80 | 0.79 |
| | BioClinical BERT | 0.78 | 0.82 | 0.74 |
| | GPT-4o | 0.73 | 0.81 | 0.77 |
| | LLAMA 3.2 | 0.84 | 0.89 | 0.79 |
| | MISTRAL7B | 0.87 | 0.90 | 0.83 |
| | RF (TF-IDF) | 0.72 | 0.85 | 0.62 |
| Bipolar | ALBERT | 0.67 | 0.77 | 0.6 |
| | MentalBERT | 0.78 | 0.86 | 0.72 |
| | BioClinical BERT | 0.77 | 0.85 | 0.70 |
| | GPT-4o | 0.73 | 0.97 | 0.59 |
| | LLAMA 3.2 | 0.83 | 0.87 | 0.80 |
| | MISTRAL7B | 0.86 | 0.88 | 0.85 |
| | RF (TF-IDF) | 0.7 | 0.91 | 0.56 |

Table 2: Classification performances during the fine-tuning process: ALBERT Model

| Mental State | F1 Score | Precision | Recall |
|---|---|---|---|
| **Suicidal/Depressed** | 0.86 | 0.83 | 0.89 |
| **Anxiety** | 0.75 | 0.76 | 0.74 |
| **Bipolar** | 0.72 | 0.78 | 0.67 |
| **Normal** | 0.82 | 0.84 | 0.80 |

Table 3: Classification performances during the fine-tuning process: MentalBERT Model

| Mental State | F1 Score | Precision | Recall |
|---|---|---|---|
| **Suicidal/Depressed** | 0.88 | 0.85 | .91 |
| **Anxiety** | 0.79 | 0.80 | 0.79 |
| **Bipolar** | 0.79 | 0.88 | 0.73 |
| **Normal** | 0.88 | 0.90 | 0.87 |

Table 4: Classification performances during the fine-tuning process: BioClinicalBERT Model

| Mental State | F1 Score | Precision | Recall |
|---|---|---|---|
| **Suicidal/Depressed** | 0.87 | 0.83 | 0.92 |
| **Anxiety** | 0.79 | 0.82 | 0.76 |
| **Bipolar** | 0.77 | 0.86 | 0.70 |
| **Normal** | 0.85 | 0.88 | 0.82 |

Table 5: Classification performances during the fine-tuning process: LLAMA Model

| Mental State | F1 Score | Precision | Recall |
|---|---|---|---|
| **Suicidal/Depressed** | 0.90 | 0.87 | 0.93 |
| **Anxiety** | 0.84 | 0.89 | 0.79 |
| **Bipolar** | 0.83 | 0.87 | 0.80 |
| **Normal** | 0.92 | 0.94 | 0.90 |

Table 6: Classification performances during the fine-tuning process: Mistral Model

| Mental State | F1 Score | Precision | Recall |
|---|---|---|---|
| **Suicidal/Depressed** | 0.92 | 0.90 | 0.94 |
| **Anxiety** | 0.87 | 0.90 | 0.83 |
| **Bipolar** | 0.86 | 0.88 | 0.85 |
| **Normal** | 0.95 | 0.96 | 0.94 |

Table 7.: Classification performances during the fine-tuning process: Random Forest (TF-IDF)

| Mental State | F1 Score | Precision | Recall |
|---|---|---|---|
| **Suicidal/Depressed** | .80 | .71 | .92 |
| **Anxiety** | .71 | .85 | .61 |
| **Bipolar** | .69 | .89 | .56 |
| **Normal** | .73 | .82 | .66 |

Table 8: Top Keywords for Depression/suicideWatch

| | |
|---|---|
| Random Forest with TF-IDF | I'm, life, feel, depression, anxiety, don't, just, want, I've, like, depressed, suicide, kill, now, die, anymore, time, feel like, friends, people |
| MentalBERT | anxiety, along, seroquel, confronted, investing, high, help, thought, quick, interest |
| LLAMA | surgical, incidence, prevention, pregnant, searching, scrounge, jager, life, defines, ensure, merit, peoplebad |

Table 9: Top Keywords for Anxiety

| | |
|---|---|
| Random Forest with TF-IDF | anxiety, I'm, feel, anxious, panic, dont, just, life, I've, like, attack, depression, want, know, time, help, feeling, going, feel like, really |
| MentalBERT | anxiety, Anxiety, medicated, work, face, trying, A, thought, results, key |
| LLAMA | sympathy, fake, adrenaline, prepurchase, loans, representations, depressant, wedding, mental, stupidest, mercilessly, niche |
